# Bleeding Risks Associated with Antidepressant Medications Among Acute Ischemic Stroke Patients on Anticoagulation or Dual Anti-platelet Therapy

**DOI:** 10.1101/2024.11.12.24317216

**Authors:** Kent P. Simmonds, Audrie Chavez, Aardhra M. Venkatachalam, Nneka Ifejika

## Abstract

**Background:** Selective Serotonin and Serotonin-Norepinephrine Reuptake Inhibitors (SSRI/SNRIs) can treat post-stroke depression and improve recovery but may be withheld for acute ischemic stroke (AIS) patients over bleeding risk concerns. Study objectives were: 1) Quantify the association between early SSRI/SNRI initiation and adverse bleeding events 2) Assess bleeding risk among patients receiving concurrent anticoagulants (AC) or dual antiplatelet therapy (DAPT) and 3) Evaluate if bleeding risks were specific to SSRI/SNRIs.

**Methods:** AIS patients were identified from Electronic Medical Records of 76 healthcare organizations (2004-2024). Patients were assigned to one of three groups within 3 months of the indexed stroke – (1) No antidepressant (No AD), (2) SSRI/SNRI, (3) Other AD (i.e., mirtazapine, bupropion, trazodone, or tricyclic ADs). The primary outcome was 1-year risk of a major bleeding event. Secondary outcomes included hemorrhagic stroke (HS), fall/fracture and death. Baseline differences were adjusted for using 1:1 matched propensity scores.

**Results:** 679,532 patients were included [No AD (n=612,868), SSRI/SNRI (n=40,136), Other AD (n=26,528)]. Early SSRI/SNRIs use (vs. No ADs) was not associated with an increased risk of a major bleed among all patients (n=36,838 pairs) or patients on anticoagulants (n=7,942 pairs). Concurrent use of SSRI/SNRIs and DAPT was associated with an 11% increased risk of a major bleed (RR: 1.11, 95% CI: 1.00 -1.24, n=7,536 pairs). Bleed risks were higher (RR: 1.10; 95 CI% 1.05, 1.14) for use of Other AD vs. SSRI/SNRI (n=22,789 pairs).

**Discussion:** SSRI/SNRIs treat post-stroke depression, promote recovery and are generally safe, however, bleed risks among patients on current DAPT should be considered.

## Introduction

Globally, stroke is a leading cause of adult disability; survivors often experience physical and psychological sequelae, resulting in substantial functional impairments.^1^ Approximately one third of stroke survivors suffer from depression, which can increase healthcare utilization, worsen functional recovery and reduce quality of life.^2–4^ Despite effective management with medications such as selective serotonin or serotonin-norepinephrine reuptake inhibitors (SSRI/SNRIs), only 1 in 3 stroke survivors with depression receive medical treatment. ^3–7^

Systemic under treatment of post-stroke depression correlates clinically with concerns regarding bleeding risk. SSRI and SNRIs inhibit serotonin reuptake by platelets, leading to impaired platelet aggregation, decreasing clot formation.^8,9^ Clinical bleeding concerns are furthered increased during the crucial early recovery period when most functional gains are achieved. Bleeding concerns are further increased among patients started on concurrent high risk medications such as anticoagulation [i.e., coumadin, direct oral anticoagulants (DOAC)] or dual anti-platelet therapy (DAPT) including aspirin + clopidogrel for secondary prevention.^10^ To date, existing studies on early initiation of anti-depressants for stroke patients have largely focused on motor recovery rather than bleeding risks.^7,11,12^

To inform clinical practice during acute and subacute recovery from stroke, we sought to:

1) Quantify the association between early SSRI/SNRI initiation and adverse bleeding events 2) Assess bleeding risk among patients receiving concurrent anticoagulants (AC) or dual antiplatelet therapy (DAPT) and 3) Evaluate if bleeding risks were specific to SSRI/SNRIs.

## Methods

A 20-year (January 1^st^, 2004 to January 1^st^, 2024) retrospective cohort of acute first-time ischemic stroke patients was generated from the electronic medical records of 76 healthcare organizations. Patients were identified using International Classification of Disease (ICD) ninth (433.xx & 434.xx) and tenth (I63.xx) revisions, with an accuracy of ∼85%.^13^ Data were acquired from TriNetX, a large analytics network platform which provides near real-time access to patient level-data; the data were internally harmonized by the TriNetX informatic team using internally verified data maps. Additional information on TriNetX data sources as well as their harmonization process was previously published.^14^ Data were considered exempt by the Institutional Review Board as it was de-identified and did not involve human subject interaction. Privacy principles employed by TriNetX are publicly available; the data and materials used for this study can be accessed via the online TriNetX platform.^15^ Study methods and results adhere to Strengthening the Reporting of Observational Studies in Epidemiology (STROBE) guidelines.^16^

### Outcomes

The primary outcome was 1-year risk of *any* major bleeding event: intracranial, gastrointestinal, pulmonary or hypovolemic shock. Secondary outcomes included the 1-year risk of non-traumatic hemorrhagic stroke (HS), fall or hip fracture, and all-cause mortality. Patient mortality records are routinely updated through the TriNetX platform. Additional details on outcomes are shown in the supplement (Table S1).

### Exposure

The primary independent variable was the use of an oral anti-depressant medication within 90 days. Three mutually exclusive groups were formed to subcategorize the exposure. 1) “SSRI/SNRIs” included citalopram, escitalopram, paroxetine, sertraline, fluoxetine, duloxetine, venlafaxine, milnacipran, or desvenlafaxine. 2) “Other ADs” included mirtazapine, bupropion, trazodone, or tricyclic anti-depressants. “No ADs” included patients with no antidepressant use over the study period. Subsequently, patients who were restarted a prior antidepressant, started an antidepressant after 90 days, or took both SSRI/SNRIs and antidepressants from other classes were excluded.

The primary comparison was between the SSRI/SNRI vs. No AD groups. A secondary comparison between the Other ADs vs SSRI/SNRI groups was performed to determine whether bleeding risks were specific to the serotonergic effect of SSRI/SNRIs.^17^

### Study Variables

Study variables were collected in the year prior to the indexed stroke and selected *a*-priori based on existing literature and perceived clinical importance.^9,18,19^ Demographic factors included age (18-59, 60-79, >80 years), sex, and race/ethnicity (White, Black, Hispanic, Asian, Other). Clinical comorbidities included absence or presence of the following conditions: hypertension, ischemic heart disease, aortic aneurysm, other aneurysms, arterial disorders, coagulation defects, diabetes mellitus, shock, Alzheimer’s disease, epilepsy, intracranial hemorrhage, subarachnoid hemorrhage, depression, anxiety, obesity, kidney disease, neoplasms, gastritis/duodenitis, gastrointestinal hemorrhage, liver disease, liver cirrhosis, osteoporosis or fall. Prior medications included platelet aggregation inhibitors (e.g., aspirin, clopidogrel, ticagrelor) and common oral anticoagulants used for secondary stroke prevention (coumadin, apixaban, rivaroxaban, and dabigatran). When available, the most recent values recorded for systolic blood pressure (<140, 140-180, 180-220, >220 mm Hg), International Normalized Ratio (<2, 2-3, 3), Alanine transaminase (<165 or ≥165 U/L), Creatinine (<2.27 or ≥2.27 mg/dL), and Hemoglobin A1c (HgbA1c) (<5.7%, ≥5.7 to <6.5%, ≥ 6.5 to <9%, ≥9%) were included.

Creatinine and alanine transaminase cut-offs as were used by the HAS-BLED score.^18^ Variables collected during the acute hospitalization included intravenous thrombolysis with tissue plasminogen activator (t-PA) and the National Institutes of Health Stroke Scale Score; categorized as mild (<10), moderate (10-19), or severe stroke (>20).

### Statistical Analysis

For each comparison, baseline differences were adjusted for using 1:1 matched propensity scores (PS).^20^ PS were based on a multivariable logistic regression model which included the 37 baseline covariates listed above. Patients were matched 1:1 using a greedy nearest neighbor matching algorithm with a caliper width of 0.25 times the standard deviation.^21^ Group differences between matched pairs were assessed using absolute standardized differences (ASDs), with ASDs >0.1 considered clinically important.^22^ Associations were estimated from relative risks (RR) and risk differences (RDs) with corresponding 95% confidence intervals (CIs). Finally, Hazard ratios (HR) with 95% CIs were estimated from Cox proportional hazard models. Schoenfeld global test evaluated the proportional hazards assumption.^23^ Results from matched pairs are interpreted the average treatment effect among the treated (ATT); ATT= E (*Y*^1^ − *Y*^0^ ∣ *D=1*) where E is the expected outcome, Y is the binary counterfactual outcome and D (1 or 0) is antidepressant use.^20^ This counterfactual framework is analogous to that used by randomized controlled trials.^24^ Important PS assumptions include: (1) all confounders are accounted for and (2) data is missing at random.^25^ All statistical analyses were two-sided, performed within the TriNetX platform with significance set at *p* <0.05.

### Subgroup analyses

Pre-specified subgroup analysis included patients on concurrent anticoagulation or DAPT as detailed in the study variables section. Select oral anticoagulants such as Edoxaban and injectable anticoagulants such as heparin or enoxaparin were not included as they are infrequently used for long-term secondary stroke prevention.^26^ DAPT included the use of aspirin 81mg or 325mg + clopidogrel 75mg.

### Sensitivity analyses

Using equivalent methodology to the primary analysis, three sensitivity analyses were performed among the following populations. 1) Patients aged >65 years (including concurrent anticoagulation or DAPT), 2) Patients who used SSRIs (i.e., SNRIs were excluded), 3) A time-restricted analysis of between January 1^st^ 2011 to January 1^st^ 2019 which corresponds with the publication dates of the Fluoxetine for motor recovery after acute ischemic stroke (FLAME) and Fluoxetine or control under supervision (FOCUS) trials.^11,12^

## Results

The study flow diagram is shown in Figure 1. Initially, 817,277 patients aged ≥18 years with their first ischemic stroke were identified. Of these, n=612,868 patients formed the No AD group. Of the n=208,409 patients who used antidepressants, n=54,986 were excluded for starting antidepressants after 3-months and n=77,674 were excluded for restarting a prior antidepressant. Finally, n=9,085 patients with antidepressant use from >1 class we excluded leaving n=66,664 patients which were subsequently split into the SSRI/SNRI (n=40,136) and Other AD (n=26,528) groups.

**Figure 1:**
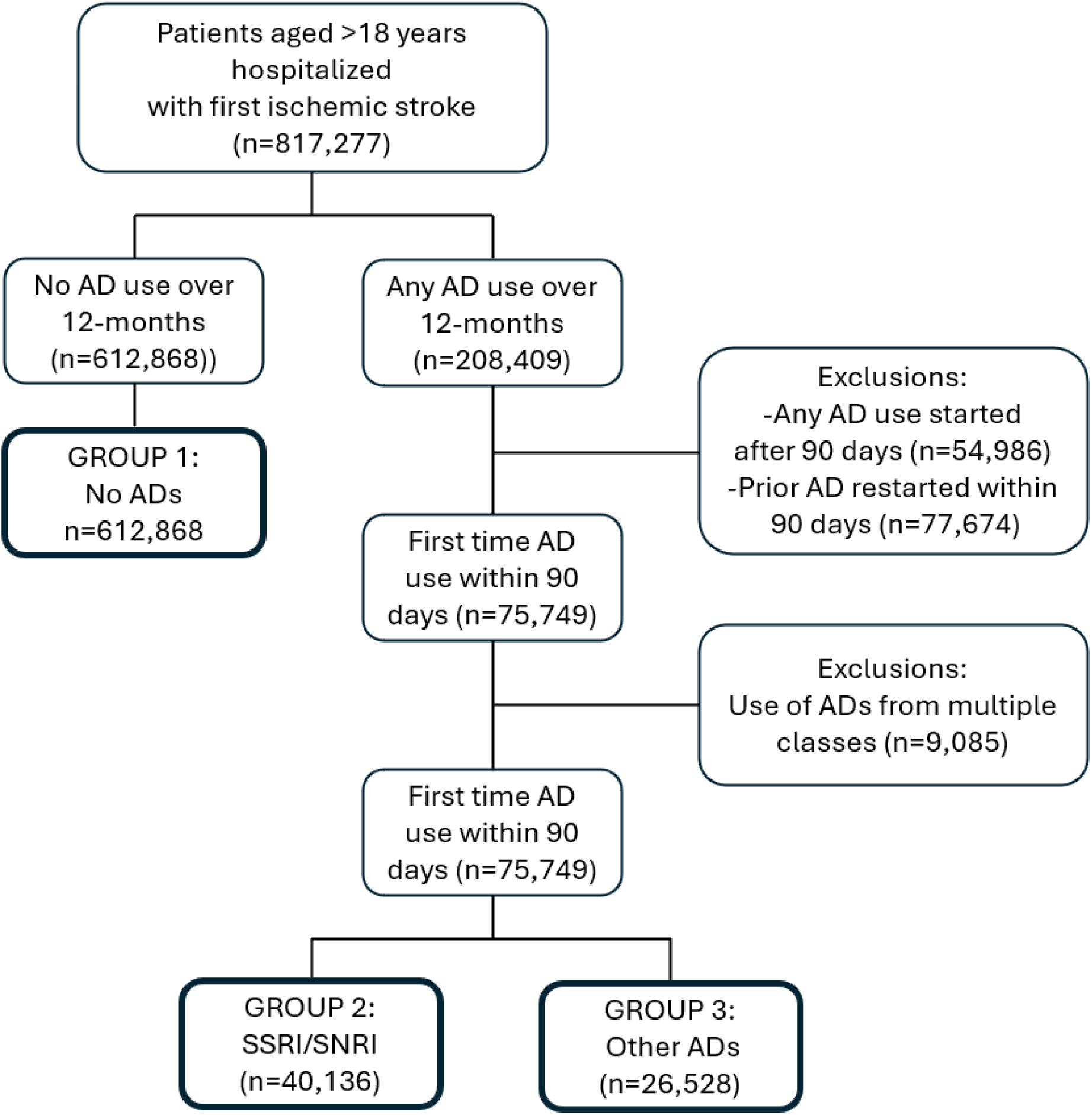
Study Flow Diagram ***Abbreviations***: AD: Anti-depressants, SSRI/SNRIs: Selective Serotonin or Serotonin Norepinephrine Reuptake Inhibitors, SSRI/SNRI group included patients who received citalopram, escitalopram, paroxetine, sertraline, fluoxetine, duloxetine, venlafaxine, milnacipran, or desvenlafaxine. Other AD group included patients who received mirtazapine, bupropion, trazodone, or tricyclic anti-depressants. No AD group included patients who did not take any of the listed AD medications over the 12-month study period

The distribution of baseline covariates is shown in Table 1. For the primary comparison, notable differences included patients who used SSRI/SNRI (vs. No AD) were more likely to be White, female, have hypertension, depression, anxiety, prior intracerebral hemorrhage, platelet aggregation inhibitor use, and receive acute stroke treatment with t-PA. Alternatively, patients not on antidepressants were more likely to be Black and have a neoplasm. For the secondary comparison, patients on Other ADs (vs. SSRI/SNRIs) were more likely to be female, have coagulation defects, prior shock, and elevated Creatine and Alanine transaminase levels.

**Table 1:**
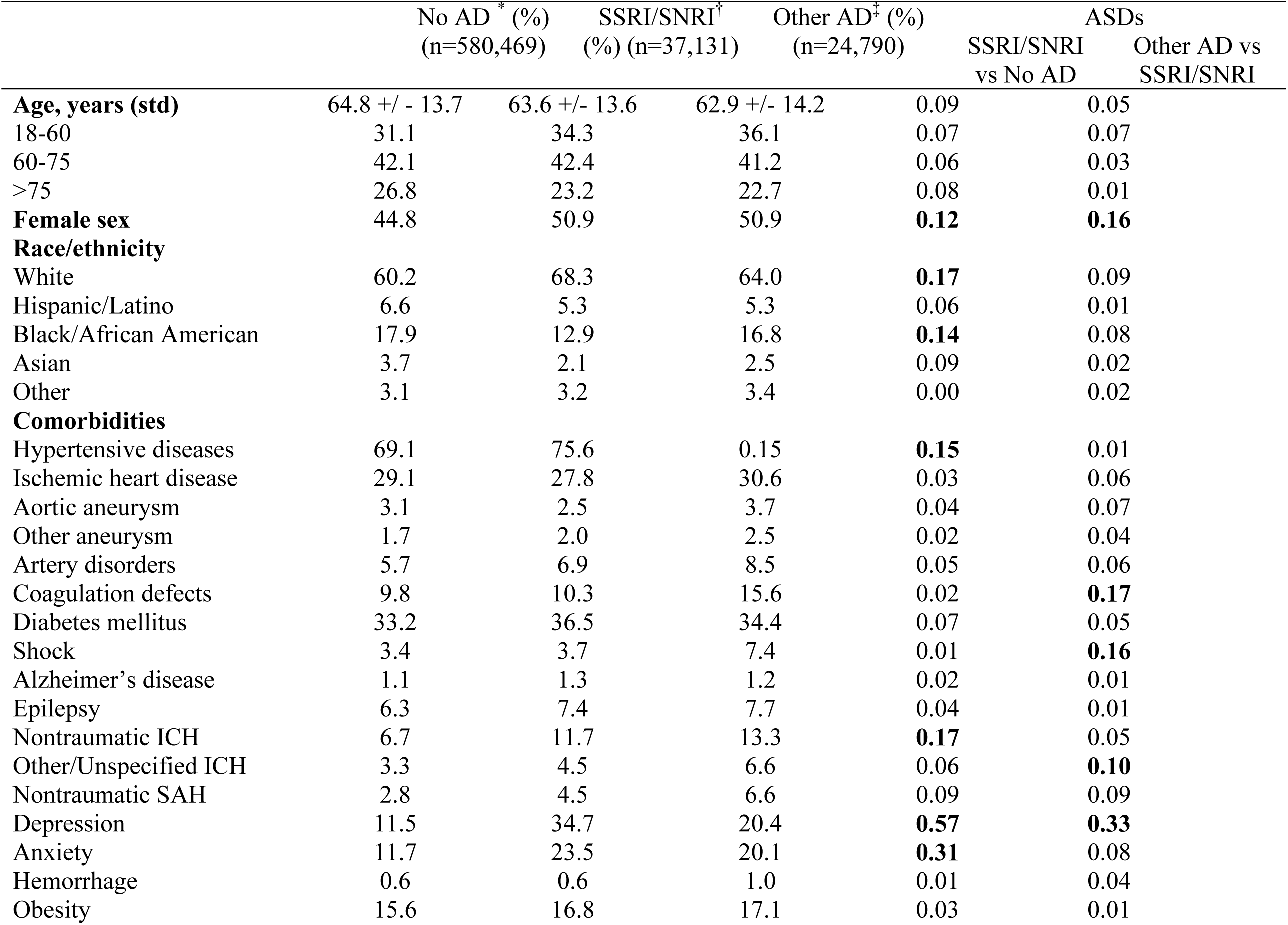
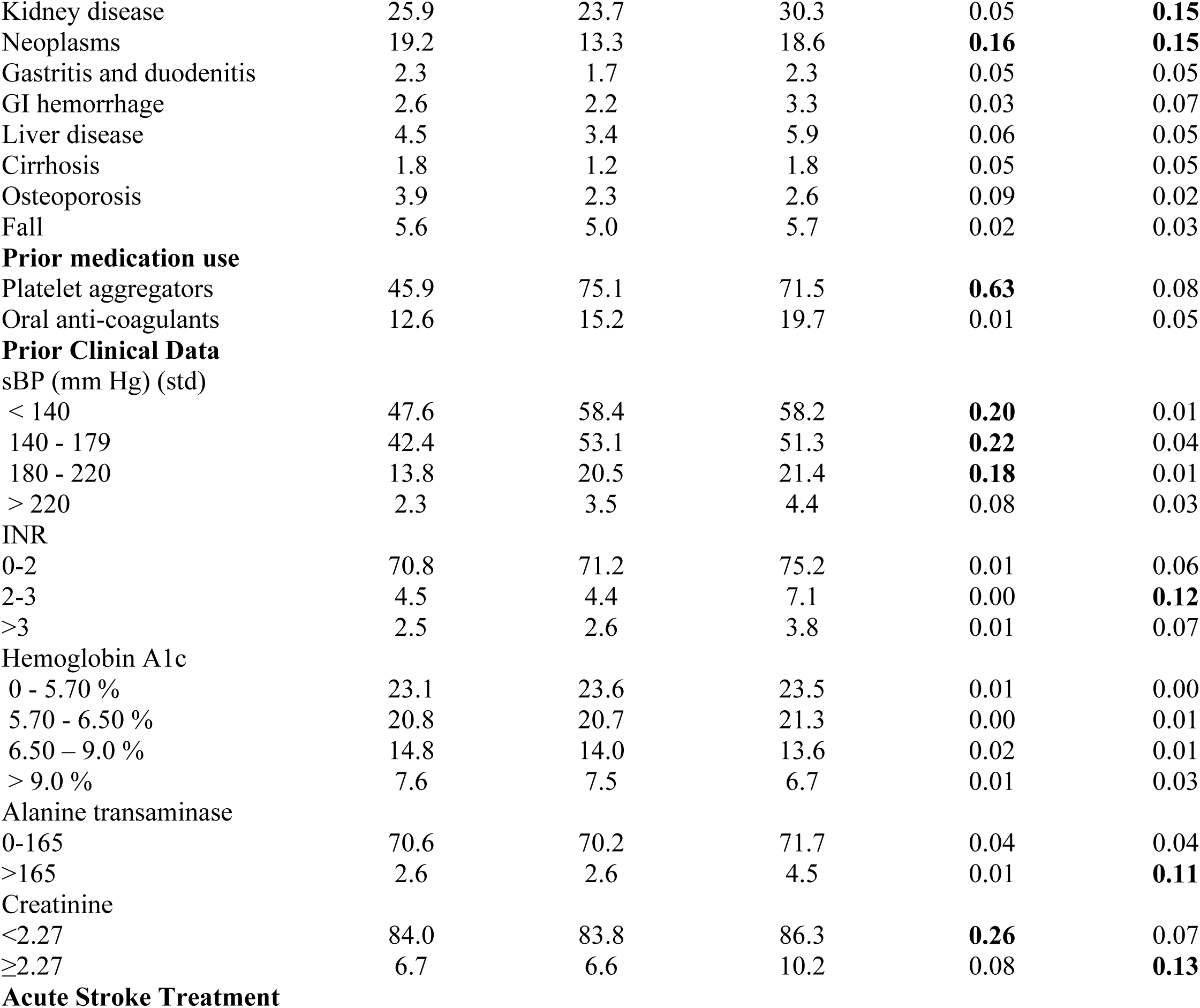
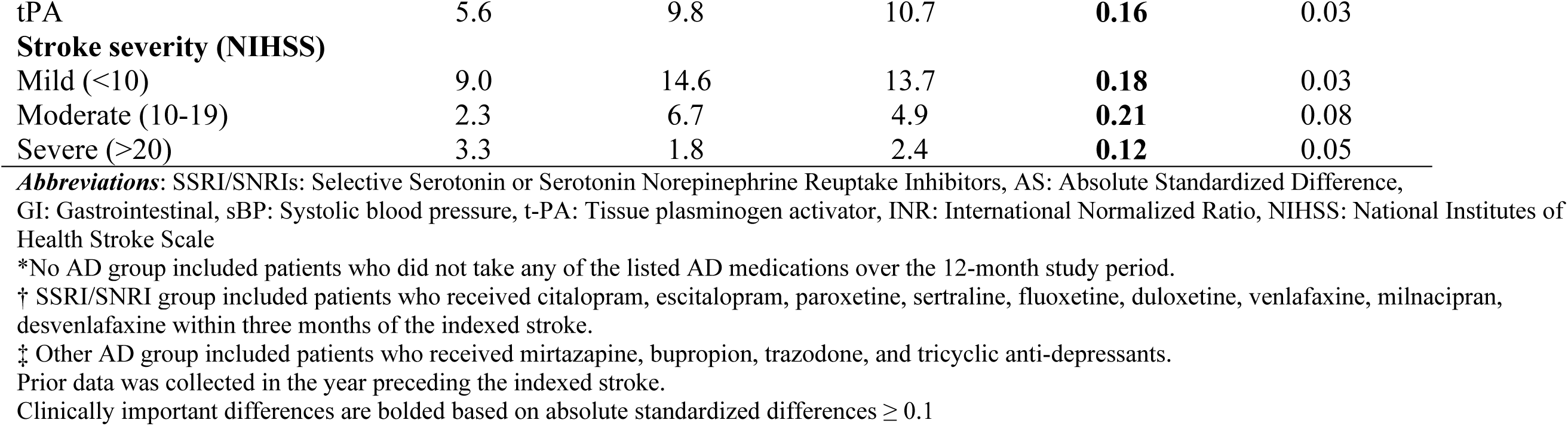
Differences in Patient-level Characteristics Among Hospitalized Acute Stroke Patients.

|  | No AD<sup>*</sup> (%)<br>(n=580,469) | SSRI/SNRI<sup>†</sup><br>(%) (n=37,131) | Other AD<sup>‡</sup> (%)<br>(n=24,790) | SSRI/SNRI<br>vs No AD | ASDs<br>Other AD vs<br>SSRI/SNRI |
| --- | --- | --- | --- | --- | --- |
| **Age, years (std)** | 64.8 +/- 13.7 | 63.6 +/- 13.6 | 62.9 +/- 14.2 | 0.09 | 0.05 |
| 18-60 | 31.1 | 34.3 | 36.1 | 0.07 | 0.07 |
| 60-75 | 42.1 | 42.4 | 41.2 | 0.06 | 0.03 |
| >75 | 26.8 | 23.2 | 22.7 | 0.08 | 0.01 |
| **Female sex** | 44.8 | 50.9 | 50.9 | **0.12** | **0.16** |
| **Race/ethnicity** |  |  |  |  |  |
| White | 60.2 | 68.3 | 64.0 | **0.17** | 0.09 |
| Hispanic/Latino | 6.6 | 5.3 | 5.3 | 0.06 | 0.01 |
| Black/African American | 17.9 | 12.9 | 16.8 | **0.14** | 0.08 |
| Asian | 3.7 | 2.1 | 2.5 | 0.09 | 0.02 |
| Other | 3.1 | 3.2 | 3.4 | 0.00 | 0.02 |
| **Comorbidities** |  |  |  |  |  |
| Hypertensive diseases | 69.1 | 75.6 | 0.15 | **0.15** | 0.01 |
| Ischemic heart disease | 29.1 | 27.8 | 30.6 | 0.03 | 0.06 |
| Aortic aneurysm | 3.1 | 2.5 | 3.7 | 0.04 | 0.07 |
| Other aneurysm | 1.7 | 2.0 | 2.5 | 0.02 | 0.04 |
| Artery disorders | 5.7 | 6.9 | 8.5 | 0.05 | 0.06 |
| Coagulation defects | 9.8 | 10.3 | 15.6 | 0.02 | **0.17** |
| Diabetes mellitus | 33.2 | 36.5 | 34.4 | 0.07 | 0.05 |
| Shock | 3.4 | 3.7 | 7.4 | 0.01 | **0.16** |
| Alzheimer's disease | 1.1 | 1.3 | 1.2 | 0.02 | 0.01 |
| Epilepsy | 6.3 | 7.4 | 7.7 | 0.04 | 0.01 |
| Nontraumatic ICH | 6.7 | 11.7 | 13.3 | **0.17** | 0.05 |
| Other/Unspecified ICH | 3.3 | 4.5 | 6.6 | 0.06 | **0.10** |
| Nontraumatic SAH | 2.8 | 4.5 | 6.6 | 0.09 | 0.09 |
| Depression | 11.5 | 34.7 | 20.4 | **0.57** | **0.33** |
| Anxiety | 11.7 | 23.5 | 20.1 | **0.31** | 0.08 |
| Hemorrhage | 0.6 | 0.6 | 1.0 | 0.01 | 0.04 |
| Obesity | 15.6 | 16.8 | 17.1 | 0.03 | 0.01 |
| Kidney disease | 25.9 | 23.7 | 30.3 | 0.05 | **0.15** |
| Neoplasms | 19.2 | 13.3 | 18.6 | **0.16** | **0.15** |
| Gastritis and duodenitis | 2.3 | 1.7 | 2.3 | 0.05 | 0.05 |
| GI hemorrhage | 2.6 | 2.2 | 3.3 | 0.03 | 0.07 |
| Liver disease | 4.5 | 3.4 | 5.9 | 0.06 | 0.05 |
| Cirrhosis | 1.8 | 1.2 | 1.8 | 0.05 | 0.05 |
| Osteoporosis | 3.9 | 2.3 | 2.6 | 0.09 | 0.02 |
| Fall | 5.6 | 5.0 | 5.7 | 0.02 | 0.03 |
| **Prior medication use** |  |  |  |  |  |
| Platelet aggregators | 45.9 | 75.1 | 71.5 | **0.63** | 0.08 |
| Oral anti-coagulants | 12.6 | 15.2 | 19.7 | 0.01 | 0.05 |
| **Prior Clinical Data** |  |  |  |  |  |
| sBP (mm Hg) (std) |  |  |  |  |  |
| < 140 | 47.6 | 58.4 | 58.2 | **0.20** | 0.01 |
| 140 - 179 | 42.4 | 53.1 | 51.3 | **0.22** | 0.04 |
| 180 - 220 | 13.8 | 20.5 | 21.4 | **0.18** | 0.01 |
| > 220 | 2.3 | 3.5 | 4.4 | 0.08 | 0.03 |
| INR |  |  |  |  |  |
| 0-2 | 70.8 | 71.2 | 75.2 | 0.01 | 0.06 |
| 2-3 | 4.5 | 4.4 | 7.1 | 0.00 | **0.12** |
| >3 | 2.5 | 2.6 | 3.8 | 0.01 | 0.07 |
| Hemoglobin A1c |  |  |  |  |  |
| 0 - 5.70 % | 23.1 | 23.6 | 23.5 | 0.01 | 0.00 |
| 5.70 - 6.50 % | 20.8 | 20.7 | 21.3 | 0.00 | 0.01 |
| 6.50 – 9.0 % | 14.8 | 14.0 | 13.6 | 0.02 | 0.01 |
| > 9.0 % | 7.6 | 7.5 | 6.7 | 0.01 | 0.03 |
| Alanine transaminase |  |  |  |  |  |
| 0-165 | 70.6 | 70.2 | 71.7 | 0.04 | 0.04 |
| >165 | 2.6 | 2.6 | 4.5 | 0.01 | **0.11** |
| Creatinine |  |  |  |  |  |
| <2.27 | 84.0 | 83.8 | 86.3 | **0.26** | 0.07 |
| ≥2.27 | 6.7 | 6.6 | 10.2 | 0.08 | **0.13** |
| **Acute Stroke Treatment** |  |  |  |  |  |
| tPA | 5.6 | 9.8 | 10.7 | **0.16** | 0.03 |
| **Stroke severity (NIHSS)** |  |  |  |  |  |
| Mild (<10) | 9.0 | 14.6 | 13.7 | **0.18** | 0.03 |
| Moderate (10-19) | 2.3 | 6.7 | 4.9 | **0.21** | 0.08 |
| Severe (>20) | 3.3 | 1.8 | 2.4 | **0.12** | 0.05 |

**Abbreviations:** SSRI/SNRIs: Selective Serotonin or Serotonin Norepinephrine Reuptake Inhibitors, AS: Absolute Standardized Difference, GI: Gastrointestinal, sBP: Systolic blood pressure, t-PA: Tissue plasminogen activator, INR: International Normalized Ratio, NIHSS: National Institutes of Health Stroke Scale
\*No AD group included patients who did not take any of the listed AD medications over the 12-month study period.
† SSRI/SNRI group included patients who received citalopram, escitalopram, paroxetine, sertraline, fluoxetine, duloxetine, venlafaxine, milnacipran, desvenlafaxine within three months of the indexed stroke.
‡ Other AD group included patients who received mirtazapine, bupropion, trazodone, and tricyclic anti-depressants.
Prior data was collected in the year preceding the indexed stroke.
Clinically important differences are bolded based on absolute standardized differences $\geq 0.1$

### Primary Comparison: SSRI/SNRI vs No Antidepressants

By one year, 13.1% of the 653,004 patients experienced a major bleed at one year. Event rates for secondary outcomes were 7.4% for HS, 5.6% for fall/fracture, and 12.8% for death.

Unadjusted results (i.e., prior to matching) results are show in Table S2. Overall, SSRI/SNRIs were associated with a statistically significant 7% increase in the risk of a major bleeding event (RR: 1.07; 95%CI: 1.04, 1.09) compared to patients not taking anti-depressants. On the absolute scale, the equivalent RD was 0.01 (95% CI: 0.01, 0.02) which translates to an overall excess risk 1 additional bleeding event per 100 patients taking SSRI/SNRIs. SSRI/SNRI use was also associated with a significant increase in the risk of HS (RR: 1.25; 95%CI: 1.21, 1.29), but a decreased risk of death (RR: 0.93; 95% CI: 0.91, 0.96). There was no significant difference in the risk of fall/fracture. Other ADs (vs. SSRI/SNRIs) were associated with an increased risk of a major bleeding event (RR: 1.33, 95% CI: 1.28, 1.37), HS (RR: 1.30, 95% CI: 1.25, 1.36) and death (RR: 1.20, 95%CI: 1.15, 1.25) (Table S3).

For the primary comparison (SSRI/SNRIs vs. No ADs), 36,838 pairs were matched with all covariates well balanced (ASDs < 0.01). SSRI/SNRIs were not associated with an increased risk of a major bleed (RR: 0.99; 95% CI: 0.96, 1.04), HS (RR: 1.03; 95%CI: 0.99, 1.08) or death (RR: 0.97; 95%CI: 0.94, 1.01). For fall/fractures, SSRI/SNRIs use were associated with statistically significant 11% reduction in risk relative to not using antidepressants (RR: 0.89; 95%CI: 0.89, 0.95). Equivalent RDs for the protective effects of SSRI/SNRIs on fall/fracture were statistically significant, however, absolute values were ≤1%. Results of aHRs estimated from the Cox proportional hazard models were largely similar (Table S4).

#### Secondary Comparison: Other Antidepressant vs SSRI/SNRIs

Results for Other ADs (vs. SSRI/SNRI) are also shown in Table 2; n=22,789 patients were matched with good covariate balance. Other ADs were associated with an increased risk of a major bleed (RR: 1.10; 95 CI% 1.05, 1.14) and HS (RR: 1.09; 95% CI: 1.05, 1.15). There was no significant difference in the risks of fall/fracture or death. Results for aHRs were largely similar (Table S4).

**Table 2:** One year risk of adverse events among acute ischemic stroke patients started on anti-depressant medications within 90 days.

|  | Primary comparison:<br>SSRI/SNRIs<sup>†</sup> vs. No ADs<sup>*</sup> (ref)<br>n=36,838 PS Matched Pairs |  | Secondary comparison:<br>Other ADs<sup>‡</sup> vs SSRI/SNRIs (ref)<br>n=22,789 PS Matched Pairs |  |
| --- | --- | --- | --- | --- |
|  | RR (95% CI) | RD (95% CI) | RR (95% CI) | RD (95% CI) |
| Any Major Bleed | 0.99 (0.96,1.04) | -0.02 (-0.52, 0.48) | **1.10 (1.05, 1.14)** | **1.52 (0.84, 2.20)** |
| Hemorrhagic Stroke | 1.03 (0.99,1.08) | 0.29 (0.12, 0.71) | **1.09 (1.04, 1.16)** | **0.99 (0.42, 1.56)** |
| Fall/Fracture | **0.89 (0.84,0.95)** | **-0.68 (-1.02, -0.33)** | 0.98 (0.91, 1.06) | -0.11 (-0.53, 0.30) |
| Death | 0.97 (0.94,1.01) | -0.35 (-0.82, 0.13) | 1.03 (0.98, 1.08) | 0.36 (-0.27, 0.98) |

**Abbreviations:** SSRI/SNRIs: Selective Serotonin or Serotonin Norepinephrine Reuptake Inhibitors,
PS: Propensity score, RR: Relative Risk, CI: Confidence Interval, RD: Risk difference
Propensity Score (PS) matched pairs: Patients were matched 1:1 based on a PS estimated from a multivariable logistic regression model with 37 covariates.
\*No AD group included patients who did not take any of the listed AD medications over the 12-month study period.
<sup>†</sup> SSRI/SNRI group included patients who received citalopram, escitalopram, paroxetine, sertraline, fluoxetine, duloxetine, venlafaxine, milnacipran, desvenlafaxine within three months of the indexed stroke.
<sup>‡</sup> Other AD group included patients who received mirtazapine, bupropion, trazodone, and tricyclic anti-depressants.

### Subgroup analysis

In the subgroup analysis n=50,712 (8%) of patients took concurrent anticoagulation, while n=85,839 (13.5%) of were on DAPT for secondary stroke prevention. PS matching resulted in n=7,942 and n=7,536 matched pairs for the anticoagulation and DAPT groups respectively with good covariate balance. For patients on concurrent anticoagulation, there were no significant differences across outcomes (Table 3). Among patients on concurrent DAPT, SSRI/SNRIs were associated with an increased risk of a major bleed (RR: 1.11; 95%CI: 1.10, 1.24) and fall/fracture (RR: 1.24; 95% CI: 1.08, 1.43) were significantly higher when compared to taking DAPT alone; with no significant difference in the risk of HS or death. Results for aHRs were largely similar (Table S5).

**Table 3:** Subgroup analysis: Comparative risks of adverse events among propensity score (PS) matched ischemic stroke patients started SSRI/SNRIs (vs. No AD) and concurrent anti-coagulation or dual anti-platelet therapy.

|  | Anti Coagulation Therapy<br>(n=7,942 PS matched pairs) |  | Dual Anti-platelet Therapy<br>(n=7,536 PS matched pairs) |  |
| --- | --- | --- | --- | --- |
|  | RR (95% CI) | RD (95% CI) | RR (95% CI) | RD (95% CI) |
| Any Major Bleed | 1.00 (0.91,1.07) | -0.14 (-1.18,0.90) | **1.11 (1.00,1.24)** | **0.88 (<0.01,1.75)** |
| Hemorrhagic Stroke | 1.04 (0.93,1.16) | 0.25 (-0.54,1.04) | 1.12 (0.965,1.31) | 0.49 (-0.15,1.13) |
| Fall/Fracture | 1.05 (0.93,1.20) | 0.29 (-0.43,1.01) | **1.24 (1.08,1.43)** | **1.06 (0.37,1.75)** |
| Death | 1.03 (0.95,1.12) | 0.38 (-0.66,1.42) | 1.04 (0.93,1.15) | 0.31 (-0.60,1.21) |

**Abbreviations:** AD: Antidepressant, SSRI/SNRIs: Selective Serotonin or Serotonin Norepinephrine Reuptake Inhibitors,
RR: Relative Risk, CI: Confidence Interval, RD: Risk difference
SSRI/SNRI group included patients who received citalopram, escitalopram, paroxetine, sertraline, fluoxetine, duloxetine, venlafaxine, milnacipran, desvenlafaxine within three months of the indexed stroke.
No AD group included patients who did not take any of the listed AD medications above over the 12-month study period.
Propensity Score (PS) matched pairs: Patients were matched 1:1 based on a PS estimated from a multivariable logistic regression model with 37 covariates

### Sensitivity analysis

Results for the first sensitivity analysis of older patients aged >65 years are shown in Table 4. Among all older AIS patients, 19,142 patients were matched, and results were similar to the primary analysis. However, contrary to the primary analysis, there was an increased risks of a major bleed (RR: 1.18; 95% CI: 1.05, 1.32), HS (RR: 1.38; 95%CI: 1.17, 1.64), and death (RR: 1.12; 95% CI: 1.01, 1.22) among older patients on concurrent SSRI/SNRIs and anticoagulants. Risks for remained elevated among older patients on concurrent SSRI/SNRIs and DAPT with respect to major bleed (RR: 1.25; 95%CI: 1.09, 1.44), HS (RR: 1.43, 95%CI: 1.16, 1.76) and fall/fracture (RR: 1.27, 95% CI: 1.07, 1.50).

**Table 4:**
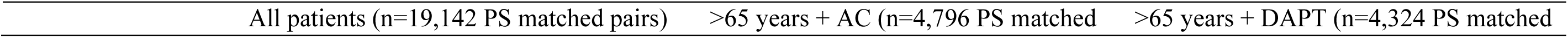
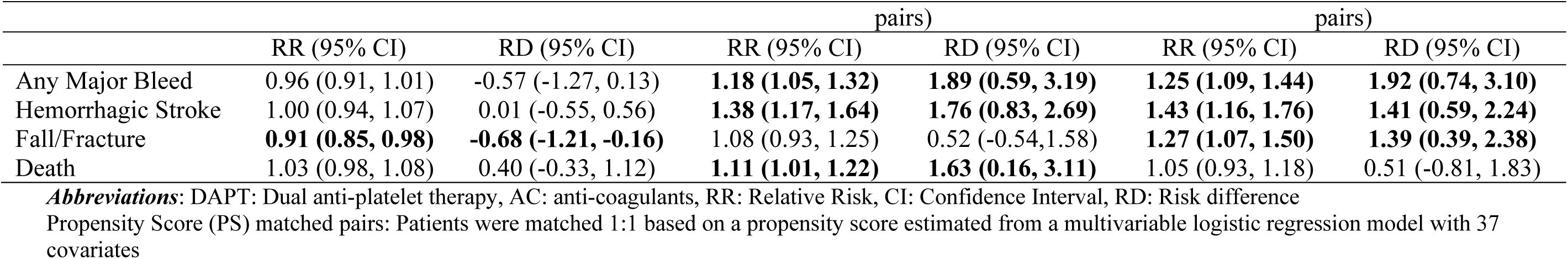
Sensitivity Analysis: Comparative risks of adverse events associated with anti-depressant mediations among acute stroke patients aged >65 years.

Results for the two other sensitivity analyses which included only patients taking SSRIs and the time-restricted analysis from January 1^st^ 2011-January 1^st^ 2019 were roughly equivalent to the main results with the exception that AD medications were associated with slight reductions in all-cause mortality Tables S6 and S7.

## Discussion

In this large multi-center study of over 650,000 patients with acute ischemic stroke, the use of SSRI/SNRIs within 3 months was *not* associated with an increased risk of a major bleeding event by one year, including among patients on concurrent anticoagulation therapy for secondary stroke prevention. Among patients on DAPT with Aspirin and Clopidogrel, SSRI/SNRI use within 3 months of ischemic stroke was associated with increased risks of bleeding and fall/fracture. Our results remained robust as results for our three sensitivity analyses of patients aged >65 years, SSRIs use, and the time-restricted analysis were largely similar to the main results. One notable exception was SSRI/SNRIs being associated with increased bleed risks among older patients aged >65 years on concurrent anticoagulation therapy.

Current literature on bleed risk associated with the use of serotonergic antidepressants is challenging to summarize due to substantial heterogeneity in clinical populations, methodological differences, and variation in the classification of bleeding outcomes. Meta-analyses non-stroke specific populations have identified SSRIs as being associated with a 66% and 51% increased risk of gastrointestinal bleeds and hemorrhagic stroke, respectively.^8,9^ Among stroke populations, a recent meta-analysis of chronic stroke patients found no association between SSRI use and gastrointestinal bleeds among ten studies (RR: 0.92, 95%CI: 0.76-1.12).^7^ Specific to AIS patients, our results were similar to a large Danish medical registry study which found no association between SSRI use and increased bleeding risk at 6-months after a stroke.^27^

To our knowledge, this is the first study to assess bleeding risks of antidepressants among AIS on current DAPT.^28^ Results from existing studies have focused on cardiac patients with mixed results. A prior study of 6,874 patients started on DAPT after percutaneous coronary intervention found no increase in bleeding risk among patients taking SSRIs.^29^ However, another study of 27,058 patients who were started on DAPT after acute myocardial infarction found concurrent SSRI use increased bleeding risks (aHR: 1.57, 95% CI: 1.07-2.32).^30^

We did not find a significant association between serotonergic antidepressants and bleeding among most ischemic stroke patients on anticoagulation. Contrary to our results, a recent meta-analysis of 13 studies identified associations between bleed risk and concurrent use of anti-coagulants and SSRIs.^28^ A single study (n=233) evaluated bleed risks among AIS patients; a significant interaction between ICH and concurrent use of SSRI was identified, but unlike our study, all patients were at increased risks of bleeding due to having also received t-PA.^31^ Similarly, increased baseline bleed risks among older patients may explain why concurrent use of SSRI/SNRIs and anticoagulants was only significant in our sensitivity analysis.^32^ Among all patients, our results were consistent with a re-analysis of 1,474 patients with atrial fibrillation, started on a DOAC from the Rivaroxaban Once Daily Oral Direct Factor Xa Inhibition Compared With Vitamin K Antagonism for Prevention of Embolism and Stroke Trial in Atrial Fibrillation (ROCKET AF) trial.^19^

Use of non-serotonergic antidepressants has previously been recommended for patients at increased risks of bleeding. Thus, our results which showed Other ADs were associated with higher risks of bleeding compared to SSRI/SNRIs were intriguing, particularly given the presumed physiological effects of serotonin on platelet aggregation.^9^ Our results partially aligned with a recent meta-analysis which found mirtazapine, a tetracyclic antidepressant, to be independently associated with an 18% increased risk of gastrointestinal bleeds.^33^ Notably, our results may be subject to residual confounding from indication bias as relative bleed risks were even higher prior to adjustment via PS matching. Indication bias would occur if clinicians elected to use “Other ADs” among patients who were more prone to bleeding out of concerns over bleeding risks associated with SSRISNRIs. Studies which directly compare bleed risks between classes of anti-depressants are lacking and we believe that there remains insufficient evidence to recommend using non-serotonergic antidepressants for patients at increased risks of bleeding.^33^

For most AIS patients, our results indicate that SSRI/SNRIs were safe to use during the crucial early recovery window of 90 days, including patients on concurrent anticoagulants. This early recovery period is crucial as it represents a crucial overlap between baseline elevations in bleed risks as well as a peak incidence of depression and anxiety. ^2934–36^ Additionally, stroke recovery is a time-sensitive, non-linear process with up to 90% of recovery often experienced by 90 days. ^34–36^ During this period, stroke patients should actively engage with rehabilitation therapies, which aim to improve cognitive, physical, psychological, and social function, mitigating long-term, permanent disability. Depression and anxiety negatively affect participation in rehabilitation therapies, which limits opportunities for achieving these functional gains and risking higher levels of chronic post-stroke debility.^36,37^

Beyond treatment of depression and anxiety, SSRIs are also associated with improvements in motor function, cognition, and dependence;^7^ albeit, with mixed evidence,^11^ particularly among patients without comorbid depression or anxiety.^38^ The 2019 update to the American Heart Association Guidelines for the Early Management of Acute Ischemic Stroke rated the evidence of motor benefit from SSRIs as weak (class IIb), and thus did not recommend their use for motor recovery.^6^ Our results demonstrating significant associations between SSRI/SNRIs and adverse events among selected patient subgroups (i.e., patients on DAPT, patients aged ≥65 years on concurrent anticoagulation or DAPT) aligns with this recommendation, as SSRI/SNRIs may result in harm.^38^ However, clinicians should consider the prevalence, morbidity, and treatability of post-stroke anxiety and depression coupled with the rarity of adverse events such as bleeds, as patient’s with untreated post stroke depression have decreased functional recovery leading increased utilization of facility-level care.^39,40^ Thus, in potentially at-risk populations, we encourage clinicians to consider the risks and benefits of early initiation of serotonergic antidepressants on an absolute, rather than relative scale given RDs better reflect true changes in risk as major bleeds among AIS are uncommon events.

Our clinical focus on the early recovery period, resulted in the generation of inception cohorts which started at the indexed stroke. While clinically informative, a limitation of this approach is the inability to control temporality between the exposure and outcomes during the 3-month window. Although our results were not affected by censoring (i.e., aHRs from the Cox proportional hazard models were roughly equivalent to RRs), we caution that our results should be interpreted as associations rather than causation. Further, if we were to restructure our inception cohorts to begin following incident antidepressant use (rather than the indexed stroke) during the early recovery period, results would be subject to immortal time bias – the error in estimating associations between exposures and outcomes that results from misclassification of time intervals.^41^

### Strengths and limitations

This study has several strengths. First, our results have strong generalizability due to the sample size and inclusion of patients from many healthcare organizations. Second, PS matches provided excellent balance across all demographic and clinical factors. Third, the pre-specified subgroup analysis among patients on concurrent anticoagulation or DAPT informs clinical decision making by providing a contextual assessment of risk among populations prone to bleeding.

Results should be interpreted in the context of several limitations. First, results are subject to residual confounding; variables captured by electronic medical records are subject to error and lack granularity. Potential confounders such as the use of non-steroidal anti-inflammatory medications were not included due to concerns of misclassification bias as their over the counter availability and intermittent use makes their utilization difficult to track.^8^ Second, the exposure variable considered *any* antidepressant use, thus our results do not account for medication dose, duration, or serotonergic affinity.^17^ Third, the primary outcome of any major bleeding event was a composite outcome based on ICD codes and may lack precision. We were unable to use standardized criteria of major bleeding events such radiographic evidence or an acute drop in hemoglobin.^42^ Finally, within the TriNetX platform, we were unable to adjust for clustering of observations within hospitals.^43^

## Conclusion

In conclusion, this large multi-center study identified that for most AIS patients, early use of SSRI/SNRIs to treat post-stroke depression and promote functional recovery are safe to start during the first three months of recovery. This includes patients on concurrent anticoagulants.

However, there were increased risks of adverse events among certain subset of patients including those on DAPT and those aged >65 years on concurrent anticoagulation or DAPT. Importantly, adverse event rates were low, and these risks be weighed against the morbidity of post-stroke depression combined with the efficacy of antidepressants.

## Data Availability

The data and materials used for this study can be accessed via the online TriNetX platform

## Abbreviations

AIS: Acute Ischemic Stroke
ASD: Absolute Standard Difference
aHR: adjusted Hazard Ratio
CI: Confidence Interval
DAPT: Dual Anti-platelet Therapy
DOAC: Direct Oral Anticoagulant
HS: Hemorrhagic Stroke
ICD: International Classification of Diseases
PS: Propensity score
RD: Risk Difference
RR: Relative Risk
SNRI: Serotonin-Norepinephrine Reuptake Inhibitor
SSRI: Selective Serotonin Reuptake Inhibitor
t-PA: Tissue Plasminogen Activator

## Disclosures

All of the authors report nothing to disclose

## Notes

### Competing Interest Statement

The authors have declared no competing interest.

### Clinical Trial

This was not a clinical trial so no registration was required

### Funding Statement

No external funding support was received

### Author Declarations

Data were considered exempt by the Institutional Review Board as it was de-identified and did not involve human subject interaction

